# Associations of PFASs and Pesticides with Lung Function Changes from Adolescence to Young Adulthood in the ESPINA study

**DOI:** 10.1101/2024.10.09.24315189

**Authors:** Kayleigh Kornher, Carlos F. Gould, Jomel Meeko Manzano, Katie Baines, Georgia Kayser, Xin Tu, Jose Suarez-Torres, Danilo Martinez, Jose R Suarez-Lopez

## Abstract

Per- and polyfluoroalkyl substances (PFASs) and pesticides are ubiquitous environmental exposures with increasingly recognized adverse health outcomes; however, their impact on lung function, particularly in combination, remains poorly understood. We included 381 adolescent participants from a prospective cohort study in Ecuador who underwent measurements of serum PFAS (perfluorooctanoic acid [PFOA], perfluorooctanesulfonic acid [PFOS] and perfluorononanoic acid [PFNA]) and urinary herbicides (glyphosate, 2,4D) and fungicides (ethylene thiourea) and had spirometric measurements in either 2016 or 2022. We characterized the association between each PFAS or pesticide and each lung function measure in log-log models estimated via ordinary least squares regression. We used quantile g-computation to assess the association of the mixture of PFAS and pesticides with lung function outcomes. After accounting for multiple hypothesis testing, and in models adjusting for household income, parental education, and exposure to tobacco, we found that, individually, PFOA, glyphosate, and ETU were associated with slight increases in FEV_1_/FVC between 2016 and 2022. No other individual associations were significant. In mixtures analyses, a one quartile increase in all PFASs and pesticides simultaneously was also not associated with statistically significant changes in lung function outcomes after accounting for multiple hypothesis testing. In large part, we do not provide evidence for associations of PFAS and herbicide and fungicide pesticides with lung function among adolescents in moderate-to-high-altitude agricultural communities in Ecuador.

## INTRODUCTION

Optimal lung function is particularly critical for adolescents, as research indicates that distinct lung function trajectories become apparent during this period, and patterns established between ages 10 and 26 are strong predictors of future respiratory health.^1,2^ Identifying predictors of adolescent lung function and lung function growth during adolescence could have major implications for improving population health.^3–5^ Environmental factors, like air pollution, are known to worsen lung function and both cause and exacerbate respiratory illnesses.^6,7^ Recent evidence has indicated that exposure to chemicals like per- and polyfluoroalkyl substances (PFAS) and pesticides could affect lung function and other respiratory outcomes through similar mechanisms.^8,9^ However, the impacts of PFAS or pesticide exposures – independently and in combination – on adolescent lung function remains poorly quantified.

In large part due to their ubiquity, environmental pollutants such as PFAS and pesticides represent increasingly large risks to public health. PFAS are a class of nearly 15,000 chemicals^10^ with strong, stable molecular structure and both lipophobic and hydrophobic properties; they used in a range of consumer products such as stain-resistant fabrics, food packaging, and non-stick cookware, and in products like firefighting foams.^11,12^ However, these properties also make PFAS highly resistant to environmental degradation and capable of long-distance transport through air and water, earning them the label ‘forever chemicals.’^13^ Consequently, humans are exposed to PFAS not only through consumer products but also through dust, food, and drinking water that have been contaminated with these substances.^14^ Human exposure to PFAS has been documented across the globe^15^, including remote regions of the Arctic^16^; perfluorooctanoic acid (PFOA), perfluorooctane sulfonate (PFOS), perfluorononanoic acid (PFNA) and perfluorohexane sulfonate (PFHxS) are the most widely detected. PFAS exposure has been associated with adverse health effects including reduced immune response to vaccines,^17^ dyslipidemia,^14^ and several types of cancer.^18^

Concerns about pesticide use and exposure have increased in recent years, particularly in agricultural regions worldwide.^19^ Herbicides glyphosate and 2,4-dichlorophenoxyacetic acid (2,4-D) widely used in both agricultural and residential settings,^20,21^ leading to increased human exposures through residues on food, inhalation of airborne particles, contamination of water sources, and accumulation in household dust.^22^ Glyphosate, 2,4-D, and ethylene thiourea (a biomarker for fungicide exposure) are frequently detected, even in non-occupationally exposed populations.^23–25^ Widespread exposures could represent a public health risk given evidence of links between exposures to pesticides like glyphosate and negative health outcomes, including cancer, ^26,27^ particularly for those living near pesticide spray sites and those who experience higher levels of exposure than general populations.^28^

Certain PFAS and pesticides are endocrine-disrupting chemicals that can interfere with hormonal signaling pathways and cause oxidative stress, affecting reproductive, developmental, and immunological systems.^13,21,29,30^ These same pathways could impair respiratory health through inflammation, oxidative stress, and immune dysregulation.^8,9,29,31,32^ Animal studies support inflammatory and immunosuppressive mechanisms. ^33–35^ Epidemiological studies in children ages 0-6, 10-15, and 0-16 have further supported these mechanisms, finding that prenatal and childhood exposures to PFAS are associated with reduced lung function among asthmatic children and higher incidence of respiratory infections in early childhood.^36–38^

However, existing work directly evaluating the effects of PFAS and pesticide exposure on lung function have yielded mixed results. Some studies have found that higher PFAS exposure worsens lung function,^37,39^ while others have found null associations.^37,40–42^ For example, drawing on cross-sectional data from 765 adolescents aged 12-19 years in the United States collected from the 2007-2012 NHANES examination, Shi et al. found no statistically significant associations between PFOA, PFOS, PFNA, and PFHxS and FEV_1_, FVC, or FEF_25%-75%_. In stratified analyses, Shi et al. found that, among girls aged 12-15 years, PFNA was negatively associated with FEV_1_/FVC, but that among boys PFNA was positively associated with FEV_1_/FVC. No associations were found among adolescents aged 16-19 years.^39^ In a cross-sectional case control study of 300 non-smoking Taiwanese children aged 10-15 years, Qin et al. found negative associations of childhood exposure to PFOS, PFOA, and PFNA with lung function, but only among those with asthma.^37^ Similarly, Kung et al. reported that prenatal exposure to PFAS was associated with lower FVC, FEV_1_, and peak expiratory flow at age eight years among a study of 165 Taiwanese children.^42^ In contrast, drawing on a prospective birth cohort of 378 Norwegian children, Kvalem et al. found no association between serum PFAS measured at 10 years old and percent predicted FEV_1_ at either 10 (cross-sectional) or 16 years old (longitudinal).^41^

Pesticide exposure has also been associated with respiratory outcomes such as asthma, lower respiratory tract infections, and impaired lung function in children, but only limited evidence exists for adolescents.^43,44^ Existing studies have largely found that pesticide exposures were associated with worse respiratory outcomes, including lower lung function, in children ^43^. For example, Raanan et al. found that exposure to organophosphate pesticides – specifically diethyl and dimethyl phosphate metabolites – during childhood was associated with lower FEV_1_ and FVC among 279 7-year-olds in the CHAMACOS cohort in Salinas Valley, California.^45^ Occupational exposure to organophosphate pesticides has also been associated with decreased lung function and increased respiratory symptoms in both adolescents and adults, as shown in studies from Egypt and India.^46,47^ While some studies have examined insecticide exposure and lung function, few studies examine the relationships between herbicides or fungicides and lung function in adolescents, with most studies focusing on occupational exposures. A meta-analysis of three studies found evidence supporting a negative association between occupational fungicide exposure and FEV_1_.^48^ Overall, research on fungicides and herbicides in relation to adolescent lung function is limited, indicating a need for more studies.

The interaction of these contaminants and their combined impact on health is not well understood. Although the effects of individual chemical groups like PFASs and pesticides are increasingly recognized, the health outcomes from their mixtures, especially in vulnerable groups such as adolescents, remain poorly documented. Shi et al. found that a mixture of PFOA, PFOS, PFNA, and PFHxS was negatively associated with FEV_1_/FVC, FEV_1_, and peak expiratory flow among 12-15-year-old girls, but not boys.^39^ However, to our knowledge, there are no studies that have characterized the health effects of mixtures of PFAS with modern-use pesticides. Existing studies assessing PFAS-pesticide mixtures have studied only legacy pesticides like organochlorine pesticides which are no longer used in most of the world and focus on prenatal exposures and neonatal or maternal outcomes^49–53^. Understanding this combined effect is particularly important given that some pesticides contain PFAS, enhancing the likelihood of joint exposures to both.^54–56^

To address this research gap, we characterized the association of exposure to PFASs, pesticides, and their mixtures with lung function in adolescents and young adults in agricultural settings. Utilizing data from a prospective cohort study, we evaluated the associations between PFAS and pesticide exposures and spirometric lung function in adolescent participants in Ecuador at two different time points.

## MATERIALS AND METHODS

### Study Design and Recruitment

The study of Secondary Exposures to Pesticides among Children and Adolescents (ESPINA: Estudio de la Exposición Secundaria a Plaguicidas en Niños y Adolescentes) is a prospective cohort study established in 2008 to examine associations between pesticide exposure and children’s development in Pedro Moncayo County, Pichincha, Ecuador. Details of ESPINA have been reported previously.^57–59^ Briefly, in 2008, a total of 313 children aged 4–9 years were recruited through community announcements and from the System of Local and Community Information (SILC) in Pedro Moncayo County.^57^ Interviews were conducted to obtain information on these children’s socio-demographic characteristics, health and pesticide exposure, and baseline data were collected at in-person examination conducted in local schools.^57^ Follow-up exams took place in 2016 (N=535 adolescents aged 11-17) and 2022 (N=505 young adults aged 17 to 24). New participants were selected and invited to participate using the SILC in 2016 and 2022. Lung function was measured in a subset of 184 participants in 2016 and all participants in 2022. Out of 783 ESPINA participants examined in 2016 or 2022, 560 had at least one lung function measurement in 2016 or 2022. Out of these participants, 431 had at least one exposure of interest measured in 2016. After excluding 50 participants with poor quality lung function measurements, there were 381 participants included in these analyses (see Figure S1 for flowchart). 102 participants had lung function measured in both years.

Informed consent of adult participants and parental permission and child assent for participation of minor participants were acquired. The institutional review boards of the University of California San Diego (UCSD), Universidad San Francisco de Quito, UTE University and the Ministry of Public Health of Ecuador approved the study. This study was also endorsed by the Commonwealth of Rural Parishes of Pedro Moncayo County.

### Examination and surveys

During the July to October 2016 exam and the July-September 2022 exams, children were assessed at seven schools during the July to October 2016 exam and six schools during the July to September 2022 exams in Pedro Moncayo County during school breaks. Examiners were blinded to participants’ potential exposures. Socio-economic and demographic information was gathered through home interviews with parents and other adult residents in 2016, and from the study participants at the examination site in 2022.

*Spirometry.* Forced vital capacity (FVC), forced expiratory volume (FEV_1_), forced mid-expiratory flow (FEF_25%-75%_) and peak expiratory flow (PEF) were measured in 2016 and 2022 using the EasyOne^TM^ Diagnostic Spirometer (NDD Medical Technologies, Zurich, Switzerland) in FVC (expiratory) diagnostic mode. This spirometer meets the standards of the European Respiratory Society (ERS) and American Thoracic Society (ATS).^60^ Participants were seated and used a nose clip. Measurements were repeated up to eight times as needed to achieve three adequate tests. Quality control was based on the quality grade determined by the spirometer. A grade of A is defined as 3 acceptable tests with a difference between best FEV1 and FVC of 100ml or less (80 ml if FVC <1 L) A grade of B is defined as 3 acceptable tests in which the difference between best FEV1 and FVC is 150ml or less (100 ml if FVC <1 L). A grade of C is defined as 2 acceptable tests in which the difference between best FEV1 and FVC is 200ml or less (150 ml if FVC <1 L) A grade of D indicates that there were 2 acceptable tests but non-reproducible results, and for a grade of F there were no acceptable tests. A, B, C were considered acceptable for FVC and FEV_1_, while only A and B were considered acceptable for FEF_25%-75%_.^60^ Percent predicted for each lung function measure, with the exception of PEF, was calculated using reference equations developed by the Global Lung Institute (GLI) via an online tool.^61,62^ Percent predicted represents the percentage of expected volume compared to what is expected for a participant’s age, height, and sex and race/ethnicity where 100% is the expected, and values above or below 100% indicate above-expected or below-expected lung parameters, respectively.

*Anthropometric measures.* Children’s height and weight were measured using a height board in accordance with the World Health Organization’s recommended guidelines ^63^ and a digital scale, respectively (Tanita model 0108 MC; Tanita Corporation of America, Arlington Heights, IL, USA).

*Venipuncture.* Blood samples were collected via venipuncture of the median cubital or cephalic veins of the arm. Samples were processed on site, and extracted serum was aliquoted and frozen on dry ice in coolers until transported to Netlab in Quito, Ecuador, for storage at -70 °C. Subsequently, samples were shipped frozen (at approximately -15 °C) using a specialized courier from Quito to the University of California San Diego (UCSD), San Diego, California USA, for long-term storage at -80 °C. Samples remained frozen during transportation. Finally, samples were shipped frozen from UCSD to a lab for PFAS quantification (see below).

*Urine collection.* Pesticide and creatinine concentrations were measured in urine samples from the July–October 2016 examination, collected at participants’ homes upon awakening. Participants brought these samples to the examination site in the morning, where they were aliquoted and immediately frozen at −20 °C. At the end of the day, the samples were transported to Quito and stored at −70 °C. These samples were subsequently shipped frozen to UCSD using a specialized courier and maintained at −80 °C upon arrival for long-term storage. For quantification of urinary metabolites, the samples were then dispatched frozen from UCSD to the respective labs via overnight shipping.

*PFAS quantification.* Concentrations of 6 PFAS were measured in serum: perfluorooctanoic acid (PFOA), perfluorooctanesulfonic acid (PFOS) and perfluorononanoic acid (PFNA), perfluorohexanesulfonic acid (PFHxS), N-ethylperfluoro-1-octanesulfonamide (EtFOSA), and perfluorooctanesulfonamide (PFOSA). The quantification procedure involved protein precipitation, centrifugation, and concentration. Samples were analyzed using liquid chromatography tandem mass spectrometry (LC/MS/MS). Specifically, a 400 μL serum aliquot was mixed with a stable isotope-labeled internal standard solution and acetonitrile in microcentrifuge tubes, and centrifuged. The clear upper layer was transferred to 96-well plates for nitrogen concentration. Analysis was performed with a Shimadzu Prominence LC system connected to an AB Sciex 5500Q mass spectrometer, using electrospray ionization in multiple reaction monitoring (MRM) mode, with retention time windows. Matrix-matched calibration curves were prepared using bovine calf serum, and quantification was achieved by isotope dilution. At least two MRM transitions were monitored per analyte, with unknowns verified against retention time and ion ratios from the calibration curve. Each batch of 20 unknown samples included three quality control (QC) samples: a method blank and low and high concentration levels, prepared in bovine calf serum. If dilution was necessary for an unknown sample, an additional QC was conducted. Batches were reanalyzed if QC recovery fell outside the 70-130% range, and unresolved QC failures led to data qualifications for the affected samples. Samples were measured at the CHEAR Exposure Assessment Hub in Minnesota.

*Urinary pesticide biomarker quantification. 2,4-D* was measured at the National Center for Environmental Health, Division of Laboratory Sciences at the Centers for Disease Control and Prevention (CDC) in Atlanta, GA. The accuracy and reliability of the analytical measurements was ensured through quality control and quality assurance protocols. If the quality control did not meet statistical evaluation standards, all study samples were re-extracted. ^40,41^. Targeted organophosphate and pyrethroid metabolites were quantified using liquid chromatography coupled with tandem mass spectrometry (LC-MS) and isotope dilution ^42^. The limit of detection (LOD) was 0.15 μg/L. Ethylene thiourea (ETU), and glyphosate were measured at the Laboratory for Exposure Assessment and Development in Environmental Research at Emory University (Atlanta, GA). To minimize potential batch effects, urine samples were randomized using a Fisher-Yates shuffling algorithm prior to analysis .^64,65^ An 800 µL aliquot of each sample was spiked with isotopically labeled internal standards and mixed with hydrochloric acid. Solid-liquid extraction cartridges were utilized for sample extraction, followed by elution with dichloromethane. The eluates were collected and concentrated to dryness, then reconstituted with formic acid. Extracts were analyzed using LC-MS with electrospray ionization. Ethylene thiourea (ETU) and its labeled analogues were measured using the multiple ion monitoring mode, with concentrations determined via isotope dilution calibration. Each analytical run included the analysis of samples, calibration curves, and quality control samples prepared in blank urine. To measure glyphosate, each sample was spiked with a stable isotopically labeled analogue of glyphosate. Solid phase extraction was utilized for sample extraction, followed by elution and derivatization steps. Analysis was performed using gas chromatography-mass spectrometry (GC-MS) with electron impact ionization. Glyphosate and its labeled analogue were measured using the selected ion monitoring mode, with concentrations determined via isotope dilution calibration. The method involved analyzing calibration curves and quality control samples prepared in blank urine. Inter-laboratory reproducibility was evaluated by participating in external quality assessment programs.

*Creatinine quantification.* Urinary creatinine concentrations were determined at the Laboratory for Exposure Assessment and Development in Environmental Research at Emory University (Atlanta, GA) using liquid chromatography electrospray ionization tandem mass spectrometry. Urinary creatinine was quantified using HPLC-MS/MS with ESI. A 10 μL aliquot of urine was diluted prior to analysis.^66^ The quantification process utilized an isotope calibration method. No further sample preparation was performed prior to analysis. The limit of detection (LOD) was established at 5 mg/dL with a relative standard deviation of 5%.

*Detectability and inclusion.* We included chemicals that were detected in 30% or more of participants for this analysis, which included PFOA (97.4%) and PFOS (97.6%), glyphosate (99.2%), ETU (89.2%), 2,4-D (66.3%), and PFNA (64.5%). Observations below the LOD were imputed as LOD/√2. However, there were machine-derived values for glyphosate (12.3%) and ETU (9.7%) that were below LOD, in which case machine derived values were used instead of imputations. Two participants had 0 ng/mL values for glyphosate; to accommodate the log transformation, these values were adjusted to half of the lowest non-zero value in the data.

### Statistical Analysis

*Covariates.* In regression models, we included income and education as indicators of socioeconomic status (SES), which tends to be associated with both exposure to environmental contaminants and worsened health, including lower lung function.^67–69^ Exposure to tobacco has also been associated with both lower lung function^70^ and higher PFAS exposure.^68^ Income, education, and tobacco exposure were assessed in 2016 in interviews with participants and their parents. Parental education was measured as the average of the years of education reported by mother and father. We included an indicator variable for whether there was a smoker in the household. Where there were missing values, we imputed parental education and household income based on 2008 values where possible or based on a random normal distribution, following previous procedures.^71^ Additionally, models that included concentrations of urinary pesticide biomarkers were adjusted for urinary creatinine to account for urine dilution. We note that our primary outcome – predicted lung function values – have already been adjusted for sex, height, and age, therefore we omit these commonly-used controls from our statistical models.

*Statistical approach.* We assessed the relationships between PFASs and pesticides and lung function in three regression approaches.

First, we estimated associations between individual PFAS and pesticide exposures and lung function outcomes (all assessed in 2016), adjusting for household incomes, total parental formal educational attainment, and the presence of a smoker in the household. We conducted log-log regression estimated via ordinary least squares (OLS), where both the exposure and outcome were natural logarithmized. Percent predicted lung function outcomes below 25% were omitted from analysis due to implausibility (N=1 FVC; N=2 FEF_25%-75%_). Coefficient estimates can be interpreted as the percent change in the lung function outcome per one percent increase in the exposure. Because we assess many individual exposure-outcome relationships, we employ a Holm-Bonferroni correction to confidence intervals to account for multiple hypothesis testing for lung function outcomes within three groups: PFAS (PFOA, PFOS, and PFNA), fungicides (ETU), and herbicides (glyphosate, 2,4-D). Presented 95% confidence intervals were still calculated where alpha = 0.05, but any reported p-values account for these corrections. This same analysis was then repeated for lung function outcomes assessed in 2022.

Second, we leveraged that we had assessed lung function at two time points in two distinct analyses. In the first, we included only participants who had lung function assessed at both time points. Here, we took the difference between 2022 and 2016 lung function outcomes and conducted a cross-sectional assessment where the outcome was 2022-2016 lung function outcomes, and the exposure of interest was 2016 assessed individual PFASs or pesticides. Here, because some participants had percent predicted lung function decline between 2016 and 2022, a logarithmic transformation was not feasible, so we opted for an inverse hyperbolic sine transformation. Changes in percent predicted lung function that were larger than 50 percentage points (positive or negative) for FVC were omitted from analysis due to implausibility. Effect estimates were then translated back to their natural units to aid in interpretation. As GLI does not provide a percent predicted for PEF that would already adjust for age, height, and sex, for this model, we adjusted for the change in age, height, and sex from 2016 to 2022 rather than the raw values in 2016 or 2022. In a second model, we leveraged the assessment of 2,4-D in both 2016 and 2022. These models included fixed effects for individual and year, thus leveraging within-subject variation in both the exposure and the outcome; this was also a log-log analysis, and standard errors were clustered at the year level. In these models, we included a variable that was defined as creatinine levels in 2016 and specific gravity levels in 2022.

Third, whereas in the previously described models we assess the independent effects of each PFAS or pesticide on lung function, recent evidence suggests that it is plausible that these exposures operate as mixtures, with effects on health outcomes that change based on the concentrations of one another.^50^ To explore this hypothesis, we employed quantile g-computation to assess the association between lung function outcomes and PFAS/pesticide mixtures (via the ‘qgcomp’ package in R). Quantile g-computation estimates the effect of simultaneously increasing all exposures by one quantile.^72^ This method allows for differentiation between exposures that have negative and positive associations with the outcomes. Weights assigned to each exposure indicate its contribution to the overall negative or positive effect measure. We logarithmized and then scaled all exposure variables such that results can be interpreted as a simultaneous one quartile increase in all exposures, conditional on included covariates (household incomes, total parental formal educational attainment, and the presence of a smoker in the household). Each mixture was assessed for the lung function in 2016, in 2022, and change (2016 measures subtracted from 2022 measures). Levels of 2,4-D measured in 2022 were included in the 2022 and change models for all outcomes. Again, a Holm-Bonferroni correction was applied to p-values to account for multiple hypothesis testing. The ‘qgcomp’ package in R computes standard errors through a bootstrapping procedure (1000 resamples).

We conducted some sensitivity analyses to assess the robustness of our models. In lieu of percent predicted models that already accounted for age, sex, and height, we also used raw spirometry values for FVC, FEV_1_, FEF_25%-75%_, and FEV_1_/ FVC as outcomes and then controlled for age, sex, and height directly in the models. Similarly, in models where percent predicted lung function was the primary outcome, we included sex, height, age, and ethnicity to check for residual confounding. In this case, all participants reported being Hispanic so this variable dropped out from the regression. In both cases, where the outcome was a change from 2016 to 2022, we controlled for change in age, sex, and height from 2016 to 2022.

Analyses were conducted using R statistical software (version 4.4.0).^73^ The ‘fixest’ package was used for cross-sectional and longitudinal regressions and the ‘qgcomp’ package was used for the quantile g-computation analyses.^74,75^

## RESULTS

**Table 1** presents characteristics and exposures of all participants as measured at the 2016 exam. **Table 2** presents age, sex, height, and raw spirometry values by year of lung function evaluation. Out of 381 participants who had measures of PFASs and pesticides from the 2016 exam, 175 had lung function measures in 2016 and 308 had lung function measures in 2022, with 102 participants having measurements in both years. In 2016, participants were, on average, 14.4 years of age (standard deviation [SD] 1.8); in 2022, participants were 20.4 years of age (SD 1.8).

**Table 1:** Descriptive statistics of households and participants (2016)

|  | **N = 381** |
| --- | --- |
| **Household income (USD/month)** |  |
| Median (Q1-Q3) | 500 (370-720) |
| **Any smokers in household** |  |
| N-Miss | 4 |
| None, N (%) | 293 (76.9%) |
| **Average of Parents' Education(years)** |  |
| Mean (SD) | 8.2 (3.4) |
| **Creatinine (ng/mL)** |  |
| N-Miss | 3 |
| Median (Q1-Q3) | 87.5 (60.4-125.0) |
| **PFOA (ng/mL)** |  |
| N-Miss | 10 |
| Median (Q1-Q3) | 0.32 (0.25, 0.39) |
| > LOQ (0.1 ng/mL), N (%) | 371 (99.5%) |
| **PFOS (ng/mL)** |  |
| N-Miss | 9 |
| Median (Q1-Q3) | 0.54 (0.39, 0.74) |
| > LOQ (0.1 ng/mL), N(%) | 372 (99.7%) |
| **PFNA (ng/mL)** |  |
| N-Miss | 8 |
| Median (Q1-Q3) | 0.07 (0.07, 0.11) |
| > LOQ (0.1 ng/mL), N(%) | 111 (29.8%) |
| **ETU (ng/mL)** |  |
| N-Miss | 3 |
| Median (Q1-Q3) | 1.46 (0.79, 2.35) |
| > LOD (0.625 ng/mL), N(%) | 303 (80.2%) |
| **Glyphosate (ng/mL)** |  |
| N-Miss | 3 |
| Median (Q1-Q3) | 0.82 (0.38, 1.72) |
| > LOD (0.25 ng/mL), N (%) | 324 (85.7%) |
| **2,4-D (ng/mL)** |  |
| N-Miss | 3 |
| Median (Q1-Q3) | 0.21 (0.11, 0.35) |
| > LOD (0.15 ng/mL), N (%) | 303(80.2%) |

*LOD* = Limit of detection, *LOQ* = limit of quantitation

**Table 2.** Sex, age, height, and spirometric parameters by year of lung function evaluation.

|  | **2016 (N = 175)** | **2022 (N = 308)** |
| --- | --- | --- |
| **Sex is female, (%)** | 51% | 50% |
| **Age (years)** | 14.5 (1.8) | 20.4 (1.8) |
| **Height (cm)** | 150.3 (9.3) | 159.2 (7.9) |
| **FVC (mL)** | 3338.5 (808.8) | 4205.5 (811.6) |
| N-Missing | 9 | 0 |
| **FEV<sub>1</sub> (mL/s)** | 2871.0 (742.8) | 3658.9 (701.8) |
| N-Missing | 9 | 0 |
| **FEF<sub>25%-75%</sub> (mL/s)** | 3422.0 (1187.3) | 4414.8 (1136.1) |
| N-Missing | 12 | 81 |
| **PEF (mL/s)** | 5914.2 (1857.1) | 8172.1 (1932.3) |
| N-Missing | 9 | 0 |
| **FEV<sub>1</sub>/FVC** | 0.88 (0.06) | 0.86 (0.08) |
| N-Missing | 9 | 0 |

Values presented are count, percentage or mean (SD)

**Figure 1** presents our percent predicted lung function outcomes and exposures by year of assessment. Scatterplots that show the joint distribution of chemical exposures and lung function outcomes are shown in **Figure 2**. The average percent predicted FVC was similar in 2016 and 2022, with averages across both years of 117% (SD 14%) for FVC. From 2016 to 2022, the average percent predicted FEV1/FVC also increased from 2016 to 2022: from an average of 96% (SD 9%) to 98% (SD 8%) (T-test P-Value = 0.02). FEF_25%-75%_ significantly increased from 109% (SD 28%) to 116% (SD 27%) (T-test P-Value=0.01). As expected, raw PEF values increased alongside participants’ growth from an average of 5910 mL/s (SD 1860) to 8170 mL/s (SD 1980) in 2016 and 2022, respectively. In 2016, the median and interquartile ranges of PFOA, PFOS, and PFNA were 0.32 ng/mL (0.25-0.40), 0.54 ng/mL (0.39-0.73), and 0.13 ng/mL (0.07-0.16), respectively. In 2016, the median and interquartile ranges of ETU, Glyphosate, and 2,4-D were 2.23 ng/mL (0.79-2.35), 0.82 ng/mL (0.38-1.72), and 0.21 ng/mL (0.11-0.35), respectively. In 2022, the median and interquartile range of 2,4-D levels was 0.27 ng/mL (0.18-0.42).

**Figure 1:**
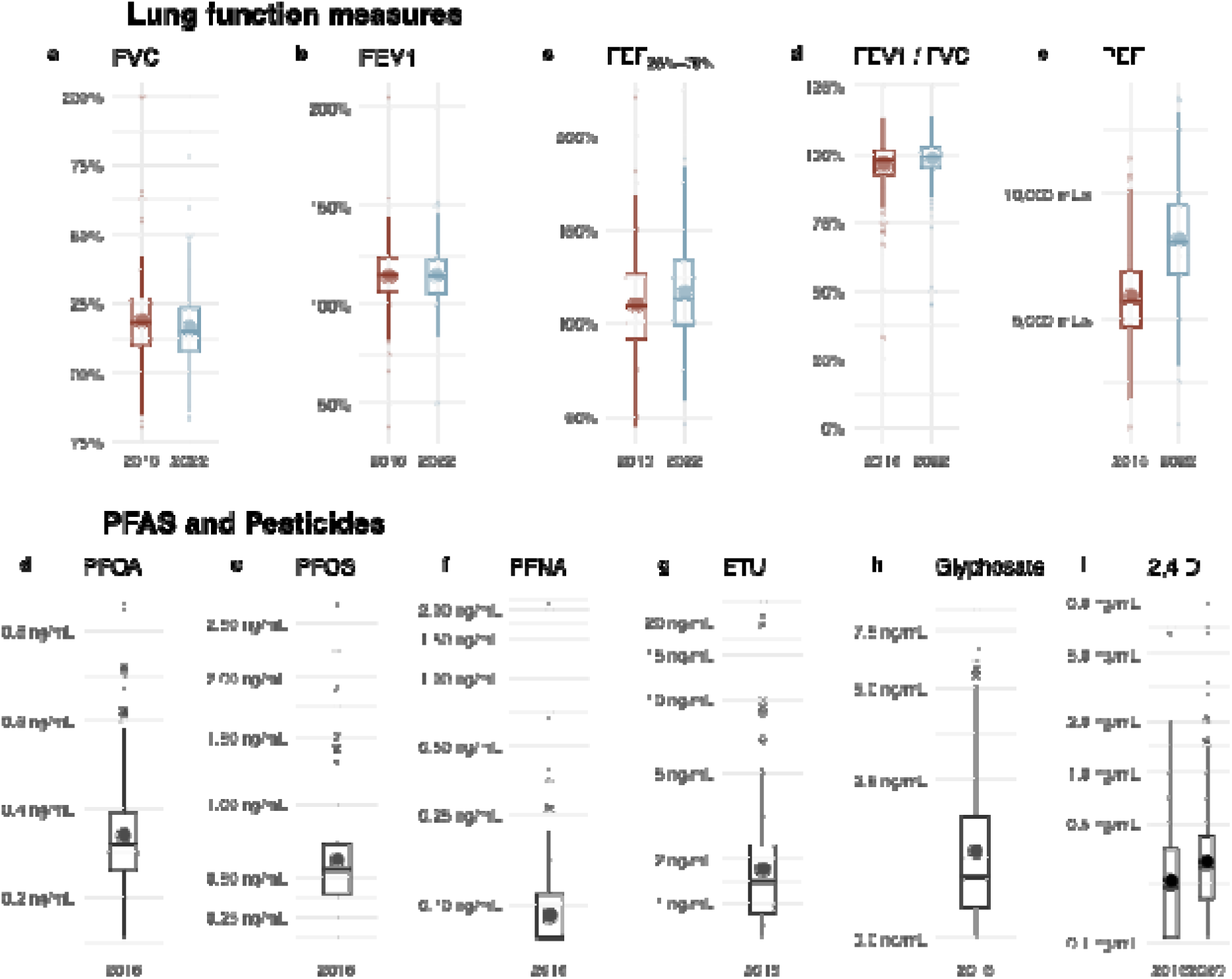
Distribution of lung function and exposure measures. Boxplots represent the median and interquartile range; whiskers extend to 2.5 times the interquartile range. Solid points represent group level means. FVC = forced vital capacity; FEF_25%-75%_ = forced mid-expiratory flow; FEV_1_ / FVC is the ratio of FEV_1_ to FVC; PEF = peak expiratory flow. PFOA = perfluorooctanoic acid; PFOS = perfluorooctanesulfonic acid; PFNA = perfluorononanoic acid; ETU = ethylene thiourea; and 2,4-D = 2,4-dichlorophenoxyacetic acid. Calculated distributions in boxplots include all data.

**Figure 2.**
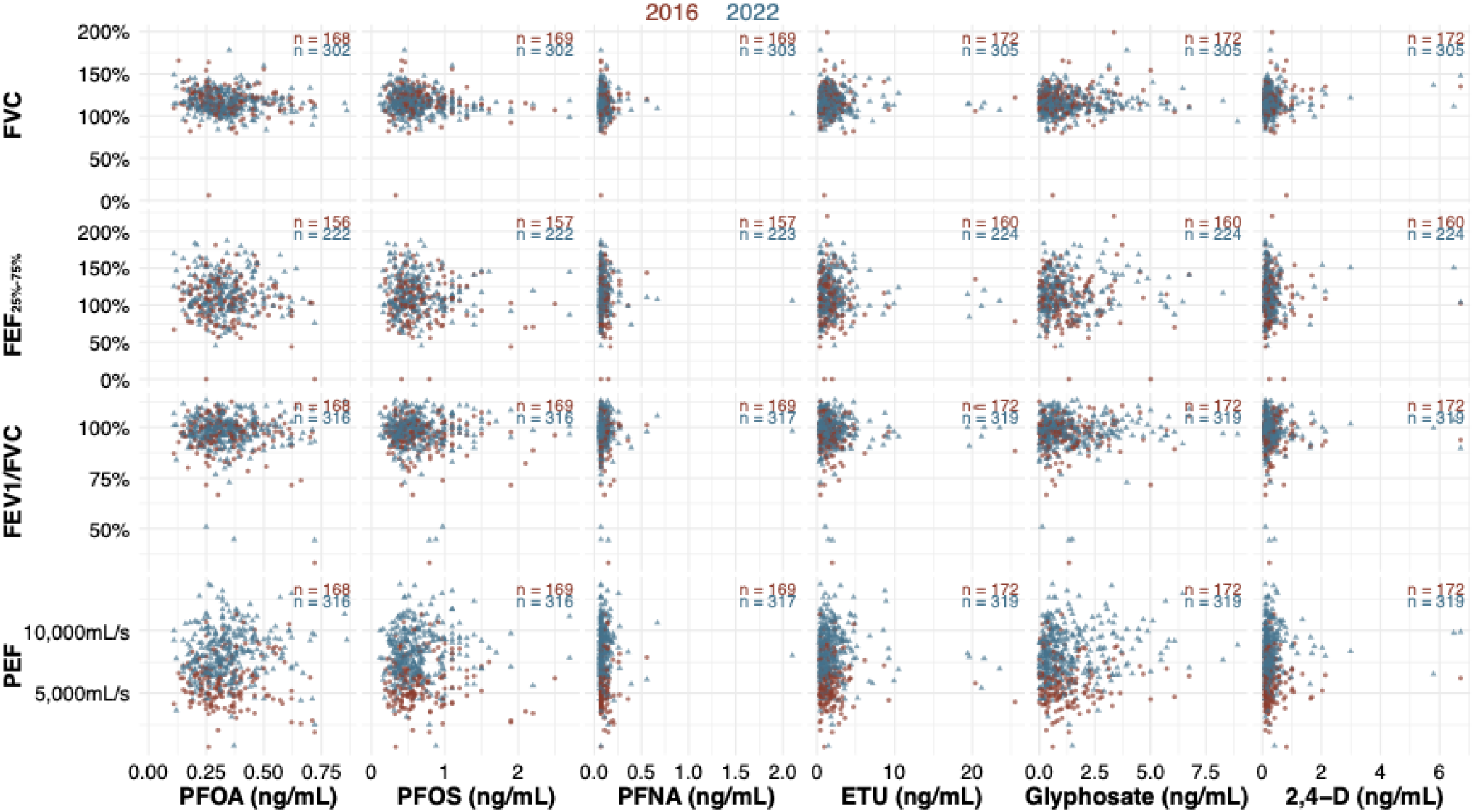
Scatter plot of chemical exposures and lung function in 2016 and 2022. Scatterplots show individual data points for 2016 chemical exposures and 2016 and 2022 lung function measures. Individual plots are annotated with sample sizes.

### Associations between PFAS and pesticides and lung function outcomes

*Cross-sectional analyses.* **Figure 3A** presents 2016 cross-sectional results from adjusted models analyzing associations of PFASs and pesticides with lung function. In adjusted models, after accounting for multiple hypothesis testing, there were no statistically significant associations between individual PFASs or pesticides and lung function outcomes assessed in 2016. Though not statistically significant, 2016 PFOA, PFOS, and PFNA exposure was negatively associated with both 2016 and 2022 percent predicted FVC. For example, a one percent increase in 2016 PFOA was associated with a 5.0% decrease (95% CI, -10.2% to 0.26%; Holm-Bonferroni p-value = 0.37) in 2016 FVC levels. There were no statistically significant associations observed for 2022 lung function outcomes (**Figure 3A**).

**Figure 3:**
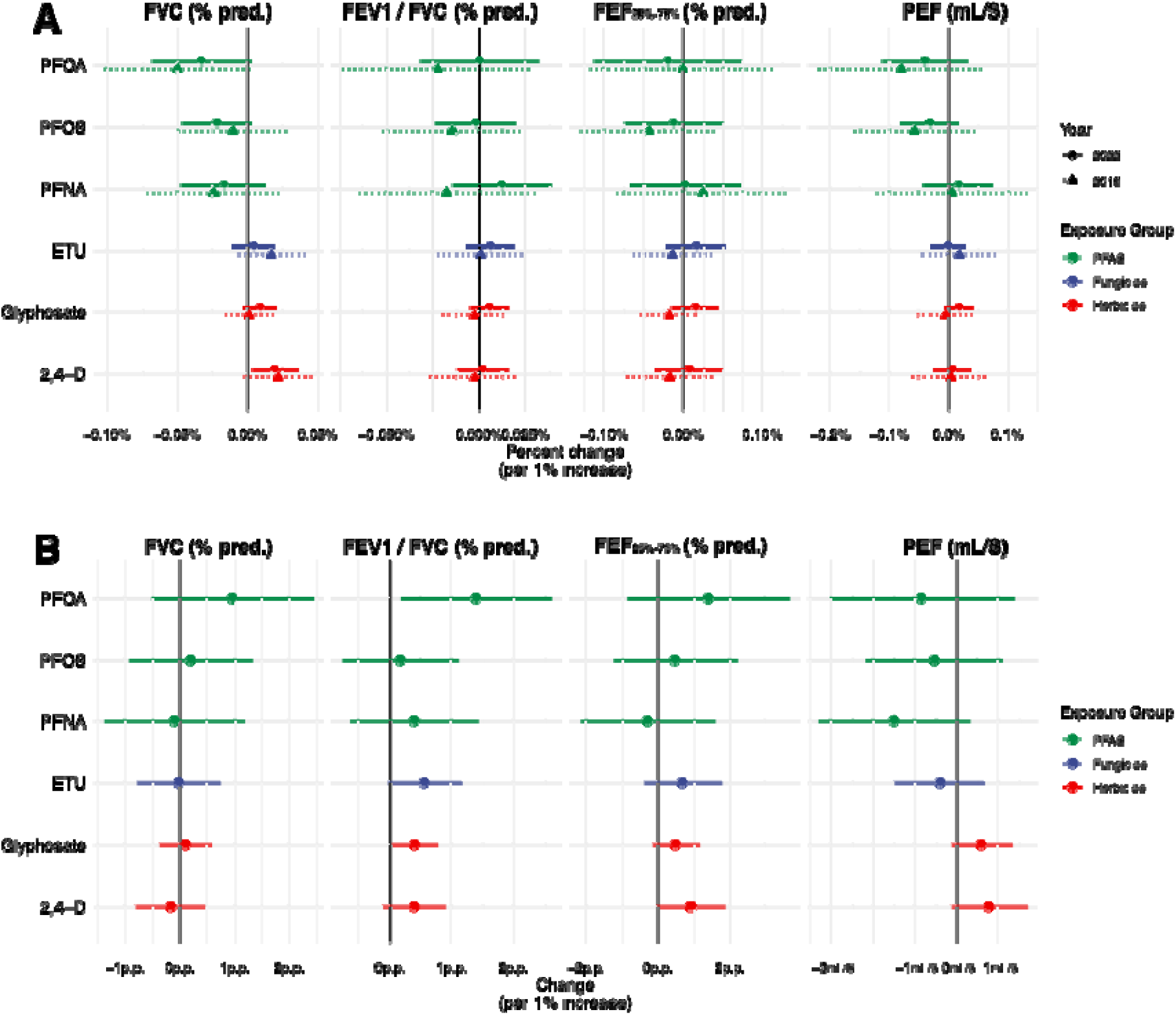
Associations of 2016 PFAS and Pesticide Exposures with Spirometric Parameters in 2016 and 2022, and Spirometric Changes (2022–2016). **(A)** displays regression coefficients (% difference in the independent variable associated [chemical exposure] with a % difference in the dependent variable [lung function]) and 95% confidence intervals from log-log models that relate 2016 exposures with 2016 spirometric parameters and, separately, log-log models that relate 2016 exposures with 2022 spirometric parameters. **(B)** displays associations between 2016 exposures and 2022-2016 spirometric parameters among participants that had lung function outcomes measured in both years. Differences were inverse hyperbolic sine transformed in analysis, and then reverse transformed into their natural units. All models adjust for parental years of education, household income, and presence of a smoker in the household. Models with urinary pesticide biomarkers as exposures also adjusted for urinary creatinine concentration. P.P = percentage point, i.e., change in percent predicted.

*Associations of 2016 PFAS and pesticides with change in spirometric parameters between 2016 and 2022.* **Supplemental Figure 1** shows individual-level changes between 2016 and 2022 in spirometric parameters and 2,4-D (the only exposure biomarker measured in both time periods). A 1% increase in PFOA was associated with a 1.9 percentage point increase (p.p.) (95% CI -0.12 to 2.8 p.p.; Holm-Bonferroni p-value = 0.62) in the change in percent predicted FEV_1_/FVC between 2016 and 2022. One percent increases in ETU (0.54 p.p.; 95% CI, -0.15 to 1.18 p.p.; Holm-Bonferroni p-value = 1) and glyphosate (0.44 p.p.; 95% CI, -0.00 to 0.88 p.p.; Holm-Bonferroni p-value = 0.40) were also associated with changes in percent predicted FEV_1_/FVC. No other meaningful associations of individual PFAS or pesticides with changes in lung function were observed (**Figure 3B**).

*Longitudinal analyses of 2,4-D and spirometric parameters, both measured in 2016 and 2022.* Leveraging both lung function and 2,4-D measurements in both 2016 and 2022 observations, we did not observe any statistically significant adjusted associations between longitudinal changes in 2,4-D and longitudinal changes in lung function outcomes (**Table 3**).

**Table 3.** Longitudinal associations between 2,4-D and spirometric parameters from 2016 to 2022.

|  | **FVC<sup>a</sup>** | **FEV<sub>1</sub> / FVC<sup>a</sup>** | **FEF<sub>25%-75%</sub><sup>a</sup>** | **PEF<sup>a,b</sup>** |
| --- | --- | --- | --- | --- |
| 2,4-D (per 1% increase)<br>(95% CI) | 1.1%<br>(-0.66%, 2.9%) | -0.80%<br>(-2.2%, 0.60%) | 2.0%<br>(-1.6%, 5.6%) | -0.81%<br>(-7.0%, 5.4%) |
| *Fixed-Effects:* |  |  |  |  |
| Participant | Yes | Yes | Yes | Yes |
| Year | Yes | Yes | Yes | Yes |
| S.E.: Clustered | Participant | Participant | Participant | Participant |
| Observations | 479 | 494 | 385 | 494 |
| R<sup>2</sup> | 0.94710 | 0.85169 | 0.90661 | 0.89283 |
| Within R<sup>2</sup> | 0.02417 | 0.00554 | 0.04852 | 0.23063 |

<sup>a</sup> Models control for creatinine levels in 2016 and specific gravity levels in 2022 as a combined variable.
<sup>b</sup> Model additionally controls for participant age and height in both 2016 and 2020.

### Mixtures

Next, we considered the associations between a simultaneous 1 quartile increase in 2016 PFOA, PFOS, PFNA, ETU, glyphosate, and 2,4-D (all logarithmized and standardized), considered as a mixture, with logarithmized lung function outcomes in 2016, 2022, and the change in lung function outcomes between 2022 and 2016 (untransformed), while adjusting for the same covariates as in our main specification. **Figure 4** presents these results as the predicted outcomes across joint exposure quartiles, relative to the second lowest quartile. There were no statistically significant associations between the chemical mixture and 2016 lung function outcomes, after accounting for multiple hypothesis testing. The mixture was non-significantly negatively associated with 2016 lung function measurements and positively associated with 2022 lung function measurements and change between 2016 and 2022. For example, a simultaneous 1 quartile increase in all PFASs and pesticides was associated with a 1.9% decrease (95% CI, - 7.0% to 3.2%; Holm-Bonferroni corrected p-value = 1) in percent predicted 2016 FEV_1_/FVC and a 2.6% increase (95% CI, 0.18% to 5.1%; Holm-Bonferroni corrected p-value = 0.44) in percent predicted 2022 FEV_1_/FVC. **Supplemental Figure 2** presents weights for each exposure component and outcome. ETU and glyphosate contributed positively to the associations most consistently, while the direction of contribution varied for other exposures. A factor distinguishing 2016 analyses from 2022 and change analyses is that 2,4-D measured in 2016 tended to contribute negatively to the associations, while 2,4-D measured in 2022 tended to contribute positively.

**Figure 4:**
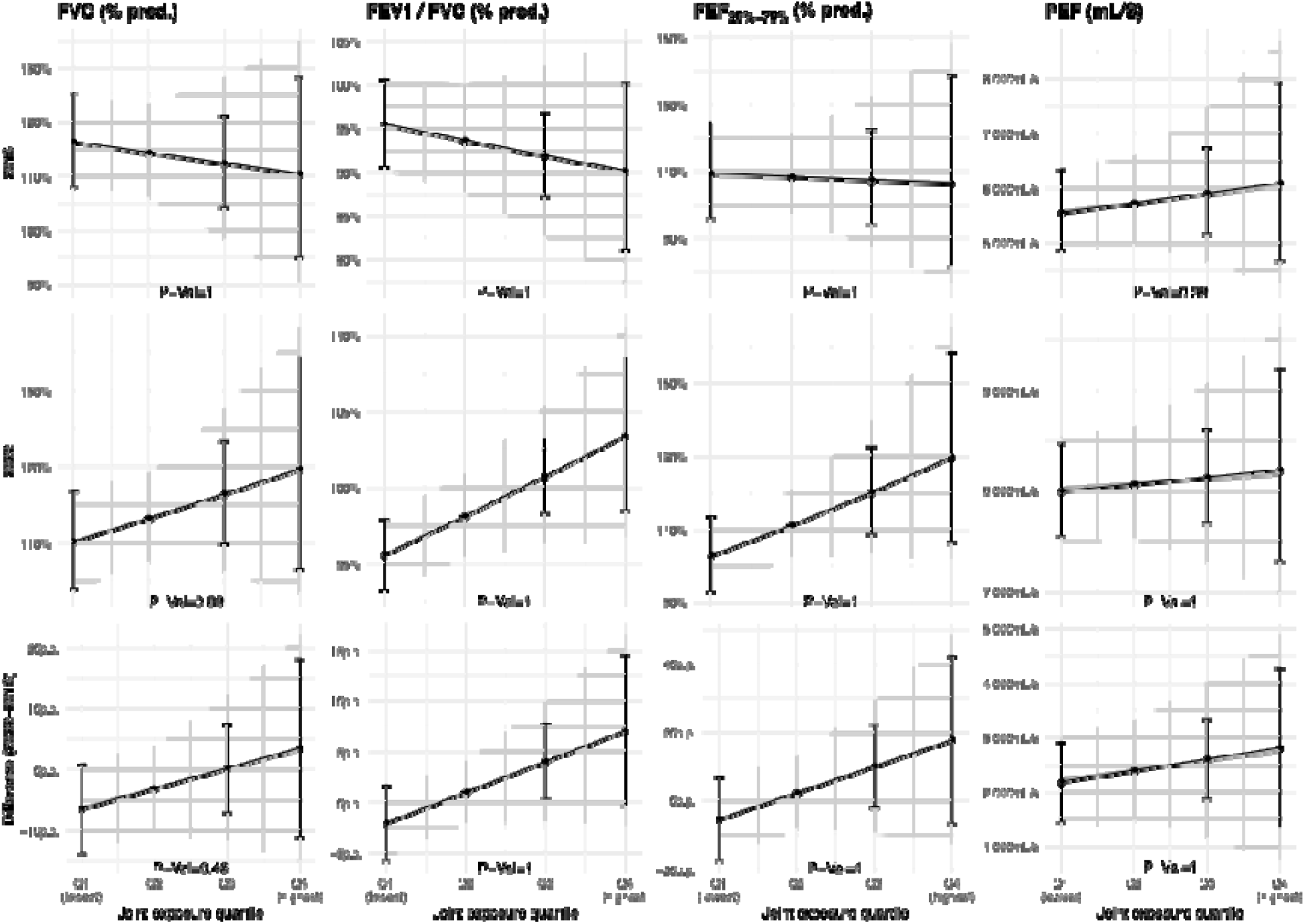
Associations between PFAS/Pesticide mixture and lung function outcomes. This figure shows the changes in percent predicted FVC, FEV_1_ / FVC, and FEF_25%-75%_ and PEF in mL/S associated with a simultaneous 1 quantile increase in PFOA, PFOS, PFNA, ETU, glyphosate, and 2,4-D, adjusted for parental years of education, household income, creatinine, and presence of a smoker in the household. Annotated p-values derived after a Holm-Bonferroni correction. p.p. = percent predicted

### Sensitivity and robustness

**Supplemental Figures 3** and **4** replicate results using raw spirometry values as opposed to percent predicted and show that results were qualitatively similar to our preferred specifications. **Supplemental Figures 5** and **6** replicate the main results where the lung function outcomes are in percent predicted, but include age, sex, and height to check for residual confounding by these variables. Results are largely unchanged.

## DISCUSSION

In a cohort of adolescents in rural Ecuador, we measured exposure to three PFASs and three pesticides through serum and urine levels and examined their association with lung function, assessed via spirometry, at two time points over a six-year period. When considered as individual exposures, we found null associations between PFOA, PFOS, PFNA, ETU, glyphosate, or 2,4-D and spirometric lung function measured in 2016 and 2022. PFOA, ETU, and glyphosate were associated with slight increases in FEV_1_/FVC between 2016 and 2022. When considered as a mixture, associations between the PFASs and pesticides included in this study and spirometric lung function in 2016 or 2022 as well as change between 2016 and 2022 were null.

Other studies have also identified null associations between individual PFAS exposure and lung function in adolescence. For example, Gaylord et al. found no association between elevated exposure to 11 PFASs, including PFOA, PFOS, and PFNA, in childhood (0-8 years) and spirometric lung function in adolescence (13-22 years).^40^ Evidence on the association between glyphosate, 2,4-D, and ETU levels and lung function in general adolescent populations is more limited, but several studies have identified decreases in lung function associated with occupational pesticide exposure in adults.^76^ The associations between many of our outcomes and exposures appear to be negative but do not reach statistical significance, which could be due to low sample size and appropriate but penalizing multiple hypothesis testing corrections. Additionally, many existing studies stratify analyses by gender or respiratory disease status, which we were unable to do due to low sample size.

Although we do not find statistically significant associations between PFAS exposures and lung function, it is plausible that PFAS exposures affect other respiratory outcomes. For example, Sevelsted et al. found that prenatal exposure to PFOS and PFOA was associated with an increased risk of non-atopic asthma by age six, while there was no significant association with atopic asthma, lung function, or atopic dermatitis.^38^ These results suggest that the relationship between PFAS exposure and lung function in children is complex and may be influenced by several factors.

Other studies have identified associations between the pesticides measured in this study and related respiratory health outcomes. Glyphosate, for example, has been associated with asthma, wheeze, and rhinitis in occupationally exposed populations.^29^ Raherison et al. found an association between urinary ETU exposure and symptoms of asthma and rhinitis in 3-10 year olds in an agricultural community in France.^77^ Islam et al identified non-significant higher odds of lower respiratory tract infections among five-year-olds exposed to ETU but no associations between 2,4-D and respiratory outcomes measured in a cohort of Costa Rican children.^44^ While lung function, our study outcome, is a valuable and objective measure of overall respiratory health, other outcomes such as asthma, wheeze, and rhinitis may also impact well-being.

Our mixtures analysis results differ somewhat from those reported in existing literature. We found that simultaneous increases in 2016 exposure to PFOA, PFOS, PFNA, ETU, glyphosate, and 2,4-D – a combination of PFASs and pesticides not previously examined in the literature – were associated with higher FEV_1_/FVC in 2022, though the relationship did not reach statistical significance after accounting for multiple hypothesis testing. Most previous individual associations identified between these exposures and lung function measures have been negative. Considering mixtures, Ye et al. identified a positive, though nonsignificant, association between exposure to a mixture of dialkyl phosphate pesticides and FVC in Canadian adolescents, findings that are similar to ours despite examining different pesticides. In the same study, these mixtures were associated with lower lung function in adults.^24^ PFAS and pesticide mixtures specifically have not been previously evaluated, but a study examining the joint effect of PUR pesticide exposure estimates (specifically carbamates, methyl bromide, and organophosphates) and air pollution exposure on lung function among asthmatic children living in an agricultural community in the San Joaquin Valley, California, USA found that higher exposure was associated with lower FVC and FEV_1_.^78^ While we adjusted for creatinine levels and several socioeconomic variables, unmeasured confounders may still influence the relationship we observed. However, our findings are consistent across models using raw spirometry values and percent predicted values adjusted for potential residual confounding.

Previous in vitro and animal studies indicate that PFASs and pesticides could reduce lung function through several mechanisms. Gestational exposure to PFOS has been shown to reduce alveolar numbers and increase lung inflammation in offspring rats, potentially inhibiting lung development.^33^ In human cells, PFAS exposure has also been linked to increased inflammation, airway hyperreactivity,^31^ and inhibition of lung surfactant function,^79^ all of which can reduce lung function. Pesticide exposure has also been linked to inflammation and oxidative stress, with potential impacts on lung function.^32,80^ In this context, it was surprising to observe positive associations with FEV_1_ of the mixtures of PFAS, herbicides and fungicides measured in our cohort. However, mixture modeling of interactions of these chemicals in relation to lung function has never been conducted; hence, replication studies are warranted.

The association of PFASs and pesticides with respiratory symptoms in early childhood identifies an additional potential mechanism by which these exposures may impact lung function. Both PFASs and pesticides – including pyrethroid insecticides and mancozeb, of which ETU is a metabolite -- have been associated with increased lower respiratory tract infections in early childhood, which are subsequently associated with lower lung function later in adolescence.^36,44,81^. This underscores the potential immunosuppressive effects of PFASs and pesticides on lung function. Moreover, it highlights the possibility of indirect pathways affecting lung function development throughout childhood and adolescence. The present study benefits from examining two time points in this critical period, allowing for the consideration of pathways that may take longer to manifest.

The population of the present study had notable differences compared to other similar studies. PFAS exposure levels were lower in our population than many other studies. For example, in the study by Shi et al, PFOA levels were approximately eight times higher, PFOS levels were over eleven times higher, and PFNA levels were approximately ten times higher than in our study^39^. Compared to the most recent levels reported in the National Health and Nutrition Examination Survey (NHANES) between 2000 and 2018 in the United States, PFOA levels were four times lower and PFOS and PFNA levels were five times lower in our study.^82^ Relatively low levels of PFASs in this Ecuadorian population may have made it more difficult to detect associations between PFAS exposure and lung function measures in this population. Pesticide levels, on the other hand, were higher than other populations examined previously. Compared to NHANES levels, 2,4-D levels measured in our study were approximately twice as high and glyphosate levels were over four times as high. However, ETU and 2,4-D levels were similar to those measured in another study on pesticide health effects in children in an agricultural community in Costa Rica.^44^ Notably, percent predicted lung function values were largely higher than expected (i.e., greater than 100% on average). Participants in the present study reside between 2,000 and 3,000 meters above sea level; people living at high altitudes have been shown to have higher lung capacity, likely as an adaptation to lower oxygen levels.^83,84^ These region-specific adaptations, coupled with lower PFAS exposures and higher herbicide and fungicide exposures than other populations could also explain the differences observed in our study with previous findings.

Our study has some limitations. First, the sample size was a limitation, particularly for the analysis of mixtures and changes in lung function, resulting in limited statistical power to detect relationships. Second, with the exception of 2,4-D, we only measured exposures in 2016, meaning we were unable to account for exposures over the six-year period. As such, the number of participants we could include in our analyses for 2022 was limited to those that had spirometric measures in both years, an important problem considering that only a subset of participants in 2016 completed spirometries. This underscores the importance of efforts to ensure consistent longitudinal follow-up, which is always a challenge in cohort studies. Third, several PFAS that we did measure were excluded from analysis due to low detectability (PFHxS, EtFOSA, PFOSA). Finally, the unique characteristics of our study population, including lower PFAS exposure, higher pesticide levels, and high-altitude residence, limit the generalizability of our findings to broader populations. However, these specificities enhance the applicability of our results to populations with similar mid-to-high altitude profiles, such as populations in the Andes, Alps, Himalayas or Rocky Mountains. Future studies, particularly those evaluating spirometric outcomes, should consider these contextual differences when designing research and comparing results and account for how PFAS and pesticide levels change over time.

## CONCLUSION

We found largely null associations between individual PFASs, herbicides, and fungicides and lung function. These findings underscore the complexity of the interactions between environmental exposures and lung function development. Future research should focus on longitudinal assessments of exposure and health outcomes, and on the effects of a broader range of chemical mixtures to better understand the multifaceted nature of environmental health risks.

## Data Availability

Private use data in this study are not publicly available.

## Acknowledgements

The ESPINA study received funding from the National Institute of Environmental Health Sciences (R01ES030378, R01ES025792, R21ES026084, CHEAR project 2018-1599, U2CES026533, U2CES026560). Dr. Kayser received funding from NIEHS K01ES031697. We also thank Fundación Cimas del Ecuador, the Parish Governments of Pedro Moncayo County, community members of Pedro Moncayo and the Education District of Pichincha-Cayambe-Pedro Moncayo counties for their support of the ESPINA study.

