## Supplemental Figure 1 for "Associations of PFASs and Pesticides with Lung Function Changes from Adolescence to Young Adulthood in the ESPINA study"

**Acknowledgements:** The ESPINA study received funding from the National Institute of Environmental Health Sciences (R01ES030378, R01ES025792, R21ES026084, CHEAR project 2018-1599, U2CES026533, U2CES026560). bvWe also thank Fundación Cimas del Ecuador, the Parish Governments of Pedro Moncayo County, community members of Pedro Moncayo and the Education District of Pichincha-Cayambe-Pedro Moncayo counties for their support of the ESPINA study.

**Supplemental Equation 1: GLI Lung Function Calculations** (From Quanjer et al., 2012)

$Predicted value=e^{a}\times H^{b}\times A^{c}\times e^{d\times group}\times e^{spline}$

Where a is the intercept, H is height in centimeters, b is the exponent for height, A is age in years and c is the exponent for age, and spline is the contribution form the age spline; group is racial group (Caucasian, African-American, South or North East Asian), and takes the value of 1 for the appropriate group and 0 for the other groups

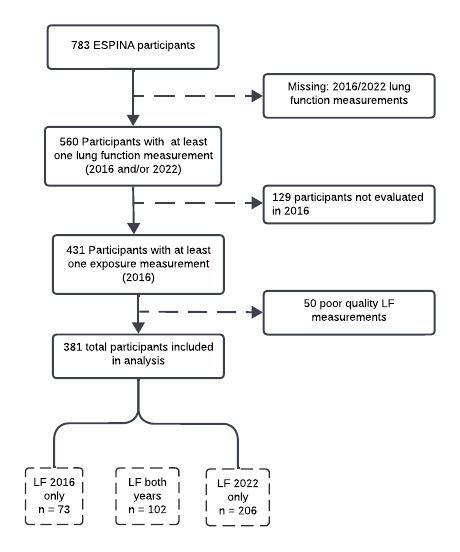

**Supplemental Figure 1: Participant Flowchart.** Out of 783 ESPINA participants, 560 had at least one lung function measurement in 2016 and/or 2022. Out of these participants, 431 had at least one exposure of interest measured in 2016. After excluding 50 participants with poor quality lung function measurements, there were 381 participants with lung function measurements in one or both years and exposure measurements from 2016.

**Supplemental Table 1: Characteristics of participants evaluated in 2016 only, 2022 only, and both years**

|  | **2016 Only (N=73)** | **2022 Only (N=206)** | **Both years --2016 (N=102)** | **Both years -- 2022 (N=102)** |
| --- | --- | --- | --- | --- |
| **Age (years)** |  |  |  |  |
| Mean (SD) | 14.6 (1.7) | 20.5 (1.8) | 14.3 (1.9) | 20.3 (1.9) |
| Range | 9.7 - 19.3 | 16.9 - 24.2 | 10.0 - 18.3 | 16.1 - 24.4 |
| **Height (cm)** |  |  |  |  |
| Mean (SD) | 149.8 (10.1) | 158.9 (8.0) | 149.7 (9.4) | 159.6 (7.8) |
| Range | 116.0 - 176.8 | 140.2 - 179.9 | 124.0 - 170.5 | 145.0 - 177.0 |
| **Weight (kg)** |  |  |  |  |
| Mean (SD) | 48.0 (9.3) | 61.7 (10.6) | 46.0 (9.4) | 60.8 (9.1) |
| Range | 28.3 - 62.4 | 34.8 - 102.6 | 26.5 - 71.0 | 38.8 - 93.3 |
| **Sex** |  |  |  |  |
| Female | 36 (49.3%) | 109 (52.9%) | 47 (46.1%) | 47 (46.1%) |
| Ma | 37 (50.7%) | 97 (47.1%) | 55 (53.9%) | 55 (53.9%) |
| **Creatinine (ng/mL)*** |  |  |  |  |
| N-Miss |  |  | 3 |  |
| Mean (SD) | 89.0 (52.1) | 107.1 (60.9) | 95.5 (58.0) |  |
| Range | 15.0 - 309.0 | 26.4 - 367.0 | 12.7 - 320.0 |  |
| **Household income* (USD/month)** |  |  |  |  |
| Mean (SD) | 576.2 (387.6) | 628.4 (522.5) | 562.1 (285.8) |  |
| Range | 100.0 - 2000.0 | 40.0 - 5000.0 | 60.0 - 1800.0 |  |
| **No. Smokers in the Home*** |  |  |  |  |
| N-Miss | 2 | 4 | 2 |  |
| 0 | 59 (83.1%) | 151 (74.8%) | 83 (83.0%) |  |
| 1 | 9 (12.7%) | 47 (23.3%) | 15 (15.0%) |  |
| 2 | 3 (4.2%) | 4 (2.0%) | 2 (2.0%) |  |
| **Average of Parents’ Education(years)*** |  |  |  |  |
| Mean (SD) | 7.9 (3.6) | 8.4 (3.2) | 8.2 (3.7) |  |
| Range | 0.0 - 17.5 | 0.0 - 17.5 | 0.0 - 19.5 |  |

*Measured in 2016 only

**Supplemental Table 2: PFAS and pesticide levels in participants with lung function evaluated in 2016 only, 2022 only, and both years**

|  | **All Participants (N=381)** | | **2016 Only (N=73)** | **2022 Only (N=206)** | **Both Years (N=102)** | |
| --- | --- | --- | --- | --- | --- | --- |
| **PFOA (ng/mL)** |  | |  |  |  | |
| N-Miss | 10 | | 4 | 3 | 3 | |
| Mean (SD) | 0.3 (0.1) | | 0.3 (0.1) | 0.3 (0.1) | 0.4 (0.1) | |
| Range | 0.1 - 0.9 | | 0.1 - 0.7 | 0.1 - 0.9 | 0.1 - 0.7 | |
| **PFOS (ng/mL)** |  | |  |  |  | |
| N-Miss | 9 | | 3 | 3 | 3 | |
| Mean (SD) | 0.6 (0.4) | | 0.6 (0.4) | 0.6 (0.4) | 0.7 (0.4) | |
| Range | 0.1 - 2.7 | | 0.2 - 2.5 | 0.1 - 2.7 | 0.2 - 2.2 | |
| **PFNA (ng/mL)** |  | |  |  |  | |
| N-Miss | 8 | | 3 | 2 | 3 | |
| Mean (SD) | 0.1 (0.1) | | 0.1 (0.0) | 0.1 (0.2) | 0.1 (0.1) | |
| Range | 0.1 - 2.1 | | 0.1 - 0.3 | 0.1 - 2.1 | 0.1 - 0.6 | |
| **ETU (ng/mL)** |  | |  |  |  | |
| N-Miss | 3 | | 0 | 0 | 3 | |
| Mean (SD) | 2.2 (3.1) | | 2.4 (3.8) | 2.5 (3.5) | 1.6 (1.2) | |
| Range | 0.3 - 25.5 | | 0.3 - 25.5 | 0.3 - 23.5 | 0.3 - 9.2 | |
| **Glyphosate (ng/mL)** |  | |  |  |  | |
| N-Miss | 3 | | 0 | 0 | 3 | |
| Mean (SD) | 1.3 (1.4) | | 1.1 (1.0) | 1.4 (1.5) | 1.3 (1.4) | |
| Range | 0.0 - 8.9 | | 0.0 - 5.0 | 0.0 - 8.9 | 0.0 - 6.7 | |
| **2,4D (ng/mL) [2022]** | 2016 | 2022 |  |  | 2016 | 2022 |
| N-Miss | 3 | 59 | 0 | 0 | 0 | 3 |
| Mean (SD) | 0.3 (0.6) | 0.5 (0.9) | 0.3 (0.2) | 0.4 (0.6) | 0.4 (0.8) | 0.5 (1.0) |
| Range | 0.1 - 6.7 | 0.1 – 9.7 | 0.1 - 1.1 | 0.1 - 6.6 | 0.1 - 6.7 | 0.1 - 9.7 |

**Supplemental Table 3: Associations between individual PFAS/pesticides and percent predicted lung function measures, including possible residual confounders**

|  | **Unadjusted** | | | **Adjusted for sex, age, height, and weight only** | | | | **Adjusted for sex, height, age, weight + other covariates** | | |
| --- | --- | --- | --- | --- | --- | --- | --- | --- | --- | --- |
|  | **FVC** | **FEV_1_** | **FEF_25%-75%_** | | **FVC** | **FEV_1_** | **FEF_25%-75%_** | **FVC** | **FEV_1_** | **FEF_25%-75%_** |
| PFOA | -12.0* (5.2) | -7.0 (7.5) | -14.2 (13.7) | | -7.9 (5.8) | -10.3 (7.6) | -20.3 (13.9) | -7.2 (5.9) | -7.0 (5.5) | -11.0 (13.9) |
| PFOS | -4.1* (1.6) | -0.5 (1.1) | -4.6 (3.9) | | -5.52 | -0.9 (1.1) | -5.3 (3.9) | -2.9. (1.7) | -0.5 (1.2) | -4.3 (4.4) |
| PFNA | -7.8** (2.7) | -0.2 (2.1) | -3.1 (5.7) | | -6.6** (2.4) | -0.7 (1.9) | -3.2 (5.6) | -6.1* (2.6) | 0.1 (1.9) | -1.0 (5.9) |
| Glyphosate | 0.7 (0.6) | 0.1 (0.3) | 0.9 (1.3) | | 0.7 (0.5) | 0.03 (0.3) | 0.9 (1.3) | 1.0 (0.6) | -0.02 (0.3) | 0.3 (1.4) |
| ETU | -0.01 (0.2) | -0.05 (0.1) | -0.4 (0.3) | | 0.1 (0.2) | -0.1 (0.1) | -0.5 (0.3) | 0.2 (0.2) | -0.1 (0.1) | -0.5 (0.4) |
| 2,4D [2016] | 2.* (1.0) | -0.01 (0.5) | 0.9 (2.0) | | 2.0. (1.0) | 0.1 (0.5) | 1.4 (2.0) | 2.1. (1.1) | 0.2 (0.5) | 1.7 (1.7) |
| 2,4D [2022] | 0.4 (0.6) | 0.4. (0.2) | 1.2 (1.0) | | 0.5 (0.5) | 0.4 (0.2) | 1.1 (1.0) | 0.4 (0.5) | 0.3 (0.3) | 1.0 (1.0) |

***=0.001, **=0.01, *=0.05, .=0.10

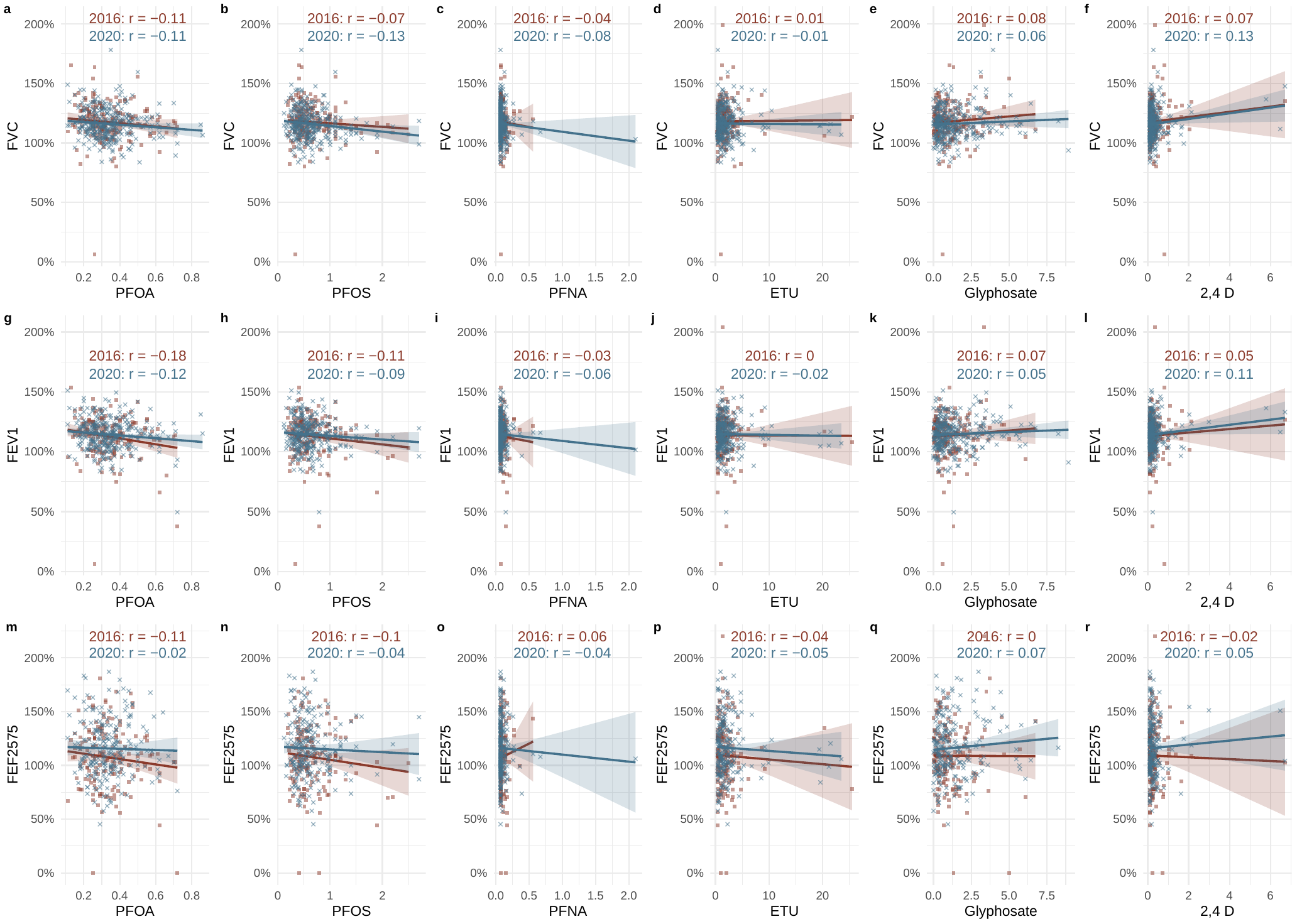

**Supplemental Figure 2. Raw correlations between exposures and outcomes by year.**

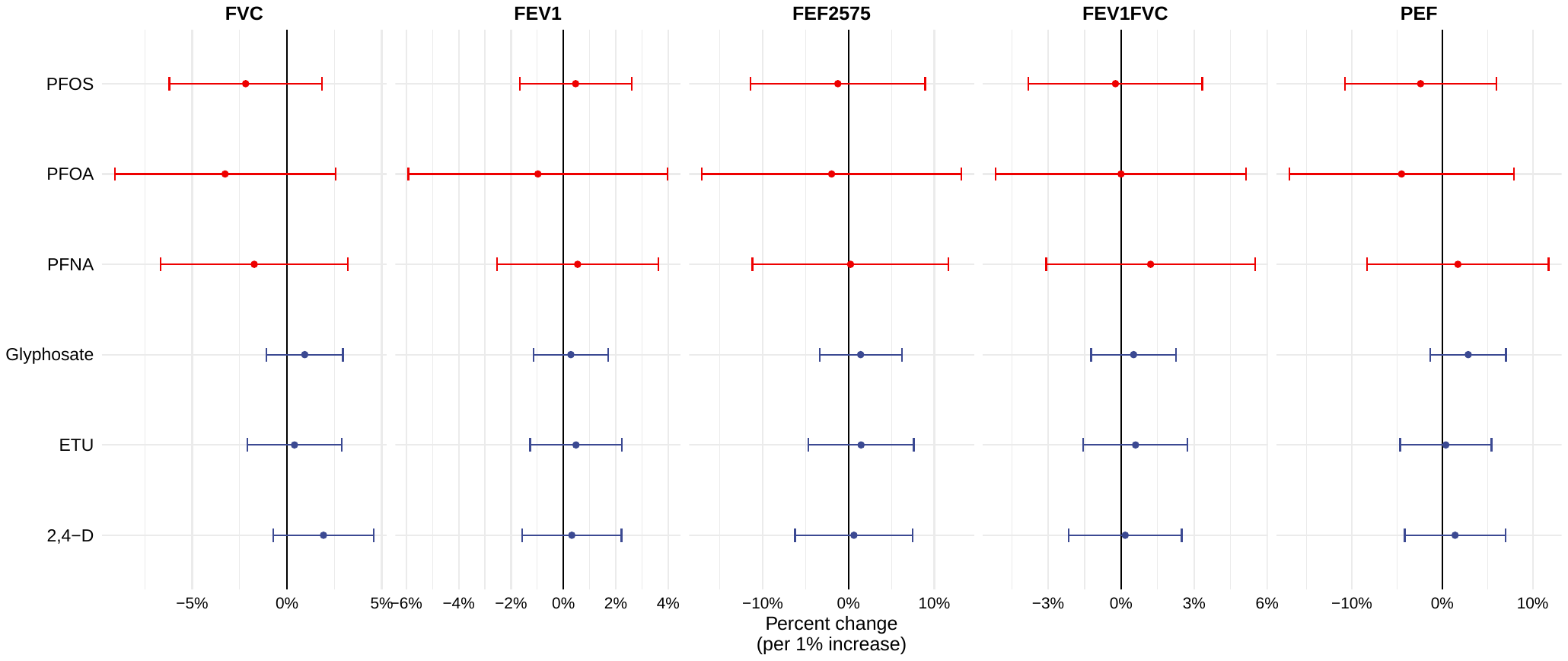

**Supplemental Figure 3: Associations between individual 2016 PFASs/pesticides and 2022 spirometry measurements, adjusted for smoking, education, income, and creatinine levels.** Regressions were log-log.

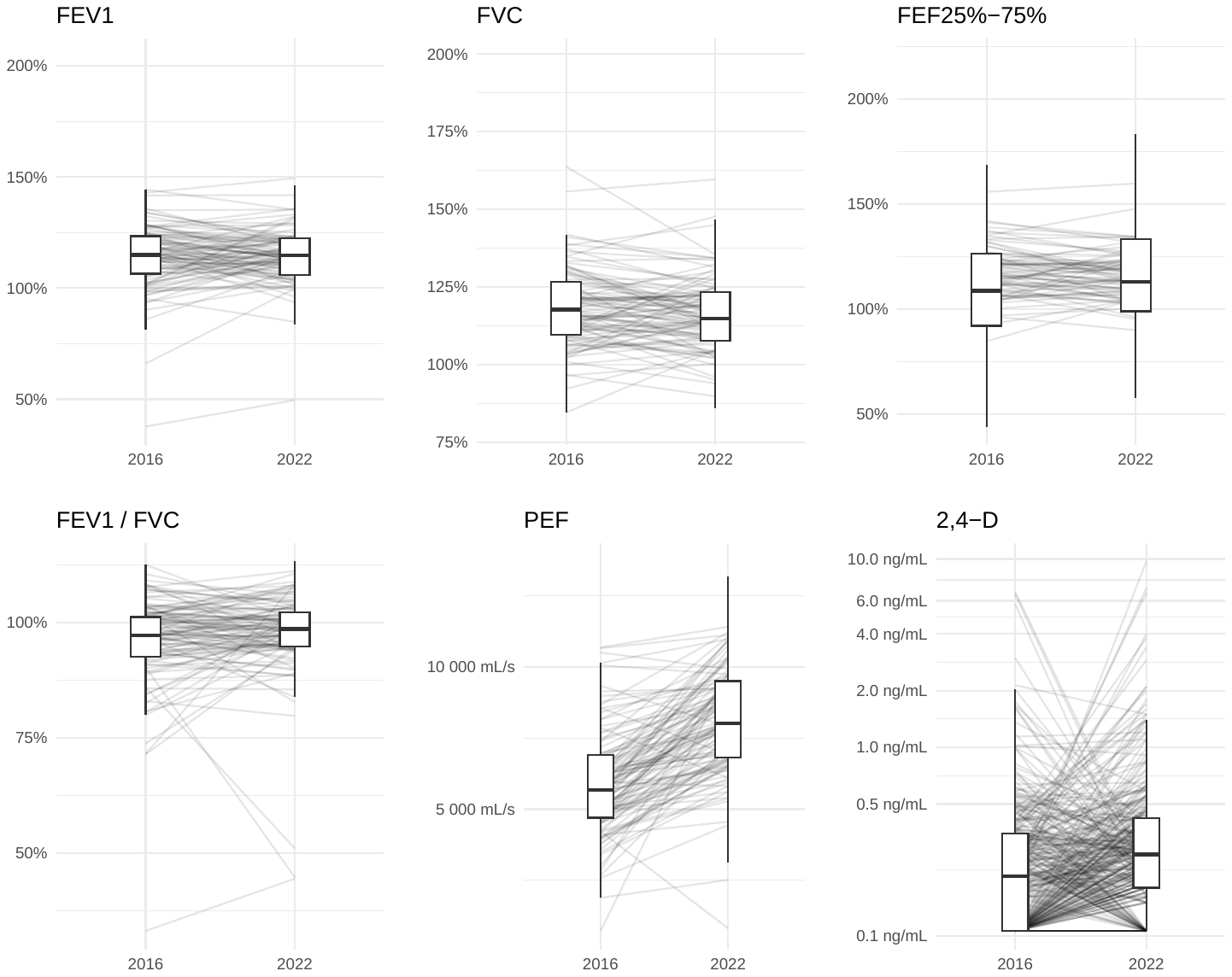

**Supplemental Figure 4. Change in spirometry and 2,4-D levels from 2016 to 2022.**

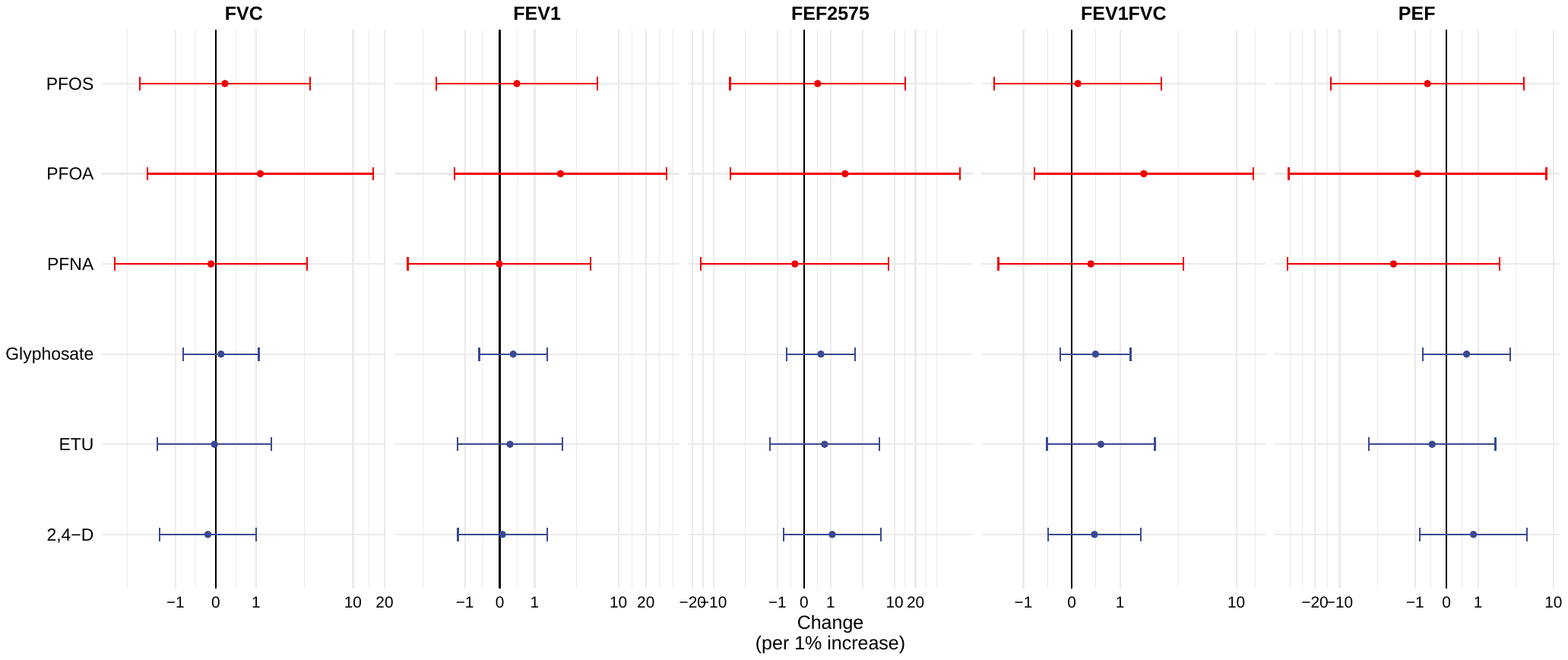

**Supplemental Figure 5: Associations between individual 2016 PFASs/pesticides and 2022-2016 spirometry measurements, adjusted for smoking, education, income, and creatinine levels.** Outcomes were inverse hyperbolic sine transformed in regressions but the coefficients have been transformed back into original units.

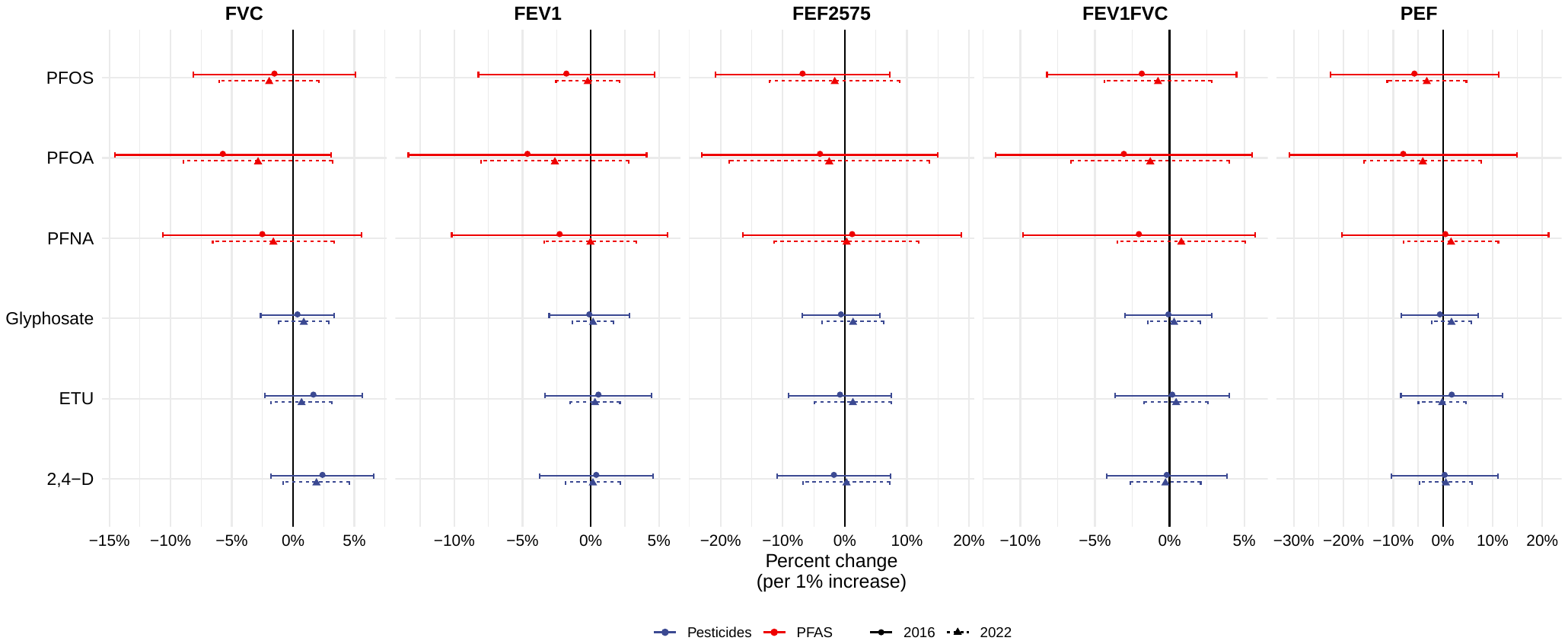

**Supplemental Figure 6: Associations between individual 2016 PFASs/pesticides and 2016 and 2022 untransformed spirometry measurements, adjusted for gender, age, and height, as well as smoking, education, income, and creatinine levels (this only for pesticides shown in blue).** Regressions were log-log.

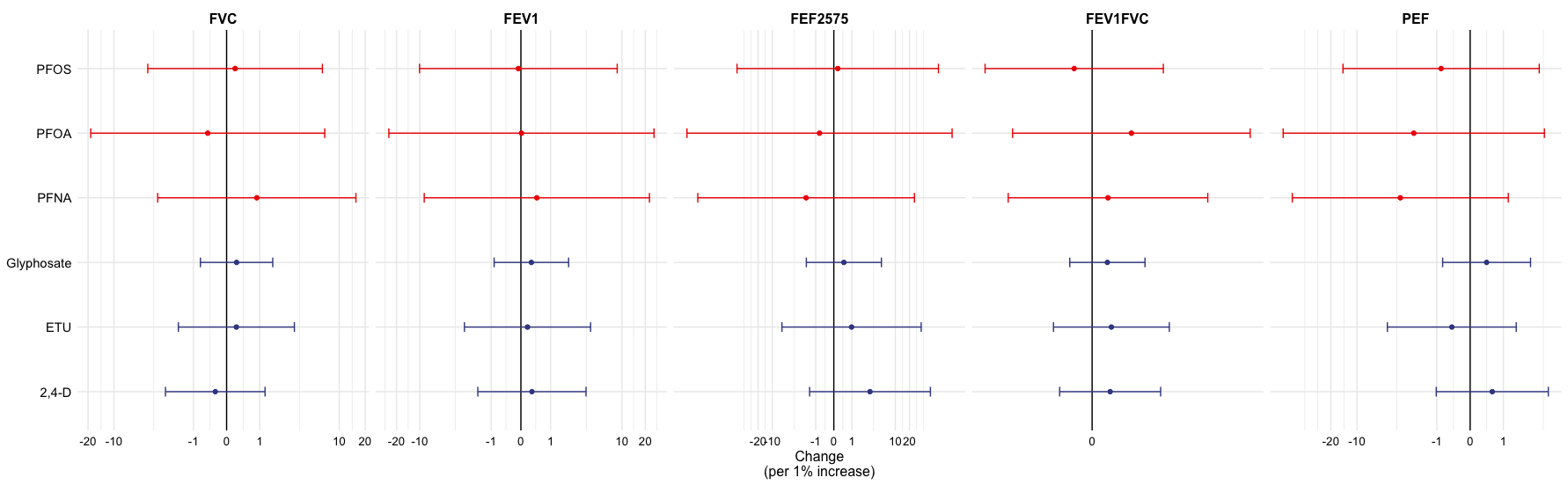

**Supplemental Figure 7: Associations between individual 2016 PFASs/pesticides and the difference between 2016 and 2022 untransformed spirometry measurements, adjusted for change in age, change in height, as well as 2016 smoking, education, income, and creatinine levels (this only for pesticides shown in blue).** Regressions were log-log.

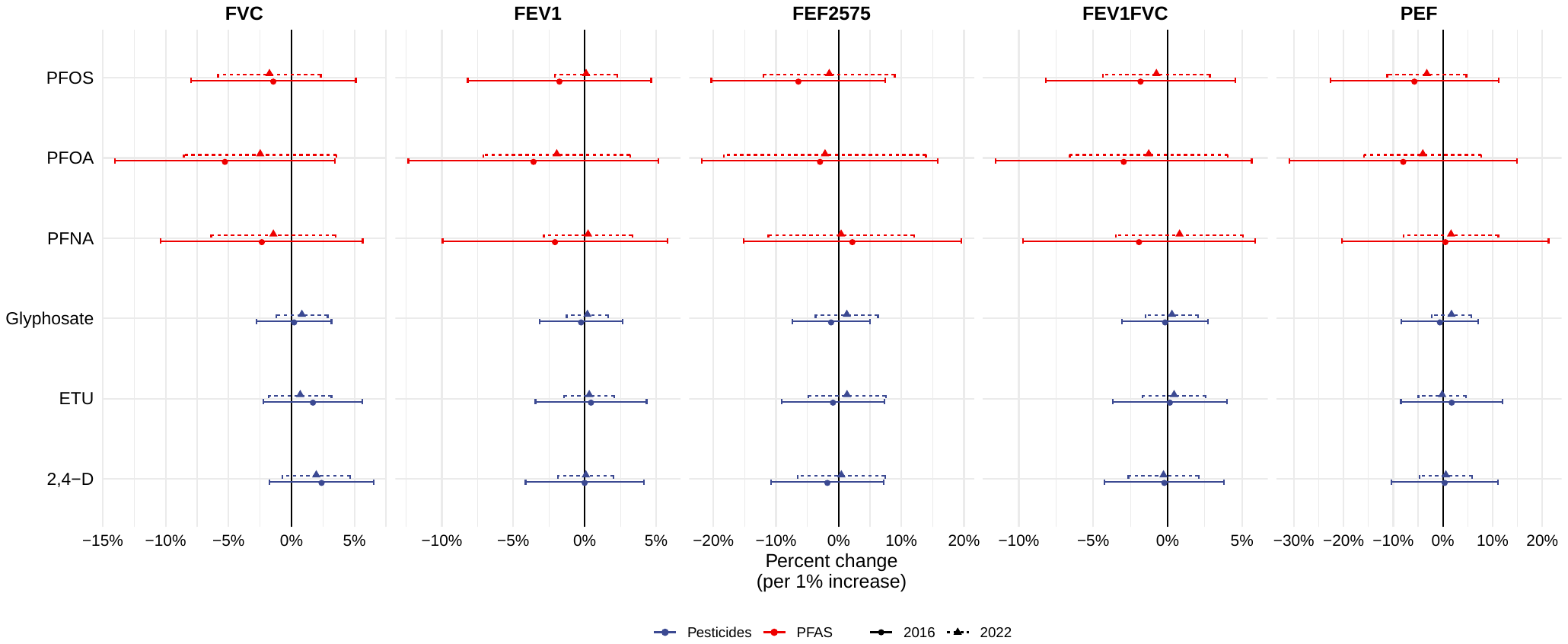

**Supplemental Figure 8: Associations between individual 2016 PFASs/pesticides and 2016 and 2022 percent predicted spirometry measurements, adjusted for gender, age, and height, as well as smoking, education, income, and creatinine levels (this only for pesticides shown in blue).** Regressions were log-log.

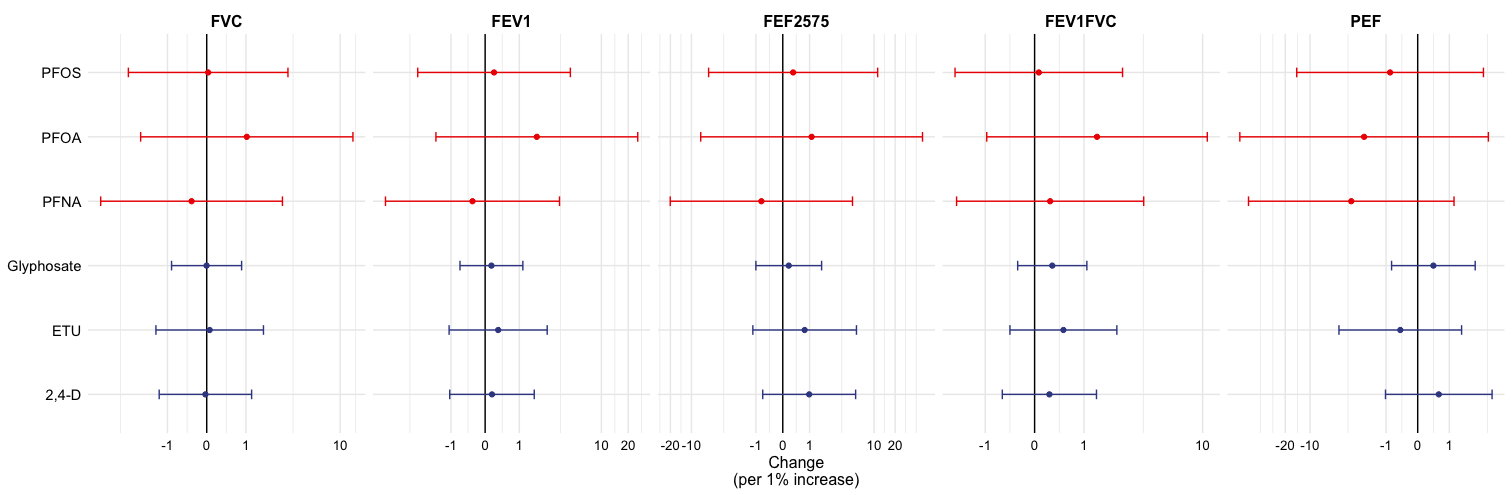

**Supplemental Figure 9. Associations between individual 2016 PFASs/pesticides and the difference between 2016 and 2022 spirometry measurements, adjusted for change in age, change in height, as well as 2016 smoking, education, income, and creatinine levels (this only for pesticides shown in blue).** Regressions were log-log.

**Supplemental Table 4. Effect of 2,4-D levels on raw lung function outcomes**

| **Dependent Var.:** | log(FVC_raw) | log(FEV1_raw) | log(FEF2575_raw) | log(FEV1FVC_raw) | log(PEF_raw) |
| --- | --- | --- | --- | --- | --- |
| **log(pest_24D)** | 0.0058 (0.0106) | 0.0023 (0.0118) | 0.0349 (0.0214) | -0.0054 (0.0075) | 0.0038 (0.0268) |
| **Fixed-Effects:** |  |  |  |  |  |
| participant | Yes | Yes | Yes | Yes | Yes |
| year | Yes | Yes | Yes | Yes | Yes |
| **S.E.: Clustered by:** | by: participant | by: participant | by: participant | by: participant | by: participant |
| **Observations** | 479 | 479 | 385 | 494 | 494 |
| **R2** | 0.98622 | 0.9771 | 0.94559 | 0.85214 | 0.89085 |
| **Within R2** | 0.84121 | 0.7613 | 0.45678 | 0.05926 | 0.21647 |

*Signif. codes: 0 '***' 0.001 '**' 0.01 '*' 0.05 '.' 0.1 ' ' 1*

*P-values corrected for multiple comparisons (alpha = 0.01).*

**Supplemental Table 5. Effect of 2,4-D levels on percent predicted lung function outcomes, adjusted for age and height**

| **Dependent Var.:** | log(FVC_pp) | log(FEV1_pp) | log(FEF2575_pp) | log(FEV1FVC_pp) | log(PEF_raw) |
| --- | --- | --- | --- | --- | --- |
| **log(pest_24D)** | 0.0107 (0.0088) | 0.0090 (0.0097) | 0.0432* (0.0190) | -0.0038 (0.0075) | 0.0038 (0.0268) |
| **Fixed-Effects:** |  |  |  |  |  |
| participant | Yes | Yes | Yes | Yes | Yes |
| year | Yes | Yes | Yes | Yes | Yes |
| **S.E.: Clustered by:** | by: participant | by: participant | by: participant | by: participant | by: participant |
| **Observations** | 479 | 479 | 385 | 494 | 494 |
| **R2** | 0. 95204 | 0. 93981 | 0. 91844 | 0. 85747 | 0.89085 |
| **Within R2** | 0. 11516 | 0. 12658 | 0. 16905 | 0. 04430 | 0.21647 |

*Signif. codes: 0 '***' 0.001 '**' 0.01 '*' 0.05 '.' 0.1 ' ' 1*

*P-values corrected for multiple comparisons (alpha = 0.01).*

**Supplemental Table 6: PFAS and Pesticide mixtures – adjusted for height, weight, sex, creatinine, no. smokers in the home, household income, and parental education.**

| **Mixture** | **LF measure** | **2016** | **2022** | **Change** |
| --- | --- | --- | --- | --- |
| PFAS and pesticides | FVC (%pred) | 1.20 (-6.64, 9.04) | 2.59 (-1.75, 6.93) | -0.89 (-10.06, 8.28) |
|  | FEV1 (%pred) | -2.06 (-7.25, 3.13) | 0.61 (-2.05, 3.27) | 4.99 (0.61, 9.37)* |
|  | FEF25%-75%(%pred) | -4.16 (-18.08, 9.77) | 4.60 (-5.52, 14.72) | 18.35 (-0.58, 37.27) |
| PFAS only | FVC(%pred) | -1.49 (-7.60, 4.61) | 0.30 (-3.38, 3.99) | -1.04 (-8.18, 6.10) |
|  | FEV1(%pred) | -1.32 (-5.34, 2.70) | 0.39 (-1.85, 2.62) | 2.30 (-1.18, 5.79) |
|  | FEF25%-75%(%pred) | -1.18 (-12.03, 9.68) | 2.02 (-6.39, 10.42) | 2.08 (-13.53, 17.70) |
| Pesticides only | FVC(%pred) | 2.60 (-1.67, 6.86) | 2.50 (0.10, 4.90)* | 0.12 (-4.77, 5.02) |
|  | FEV1(%pred) | -0.58 (-3.39, 2.24) | 0.24 (-1.24, 1.72) | 1.84 (-0.54, 4.23) |
|  | FEF25%-75%(%pred) | -3.03 (-10.54, 4.47) | 2.36 (-3.51, 8.24) | 12.65 (2.19, 23.10)* |

**Supplemental Table 7: Quantile g-computation weights**

| **Year** | **FVC** | | **FEV1** | | **FEF_25%-75%_** | |
| --- | --- | --- | --- | --- | --- | --- |
|  | PFOA (-) | 0.74 | PFOA (+) | 0.57 | PFOA (+) | 0.33 |
| 2016 | PFNA (-) | 0.14 | PFNA (-) | 0.51 | PFNA (-) | 0.36 |
|  | ETU (+) | 0.37 | ETU (-) | 0.06 | ETU (+) | 0.51 |
|  | Glyphosate (+) | 0.24 | Glyphosate (+) | 0.43 | Glyphosate (-) | 0.20 |
|  | 2,4D (-) | 0.22 | 2,4D (-) | 0.32 | 2,4D (-) | 0.45 |
|  | PFOA (-) | 0.17 | PFOA (-) | 1.0 | PFOA (+) | 0.11 |
|  | PFOS (-) | 0.65 | PFOS (+) | 0.06 | PFOS (-) | 1.0 |
| 2022 | PFNA (+) | 0.29 | PFNA (+) | 0.43 | PFNA (+) | 0.29 |
|  | ETU (-) | 0.18 | ETU (+) | 0.15 | ETU (+) | 0.20 |
|  | Glyphosate (+) | 0.37 | Glyphosate (+) | 0.08 | Glyphosate (+) | 0.17 |
|  | 2,4D (+) | 0.24 | 2,4D (+) | 0.29 | 2,4D (+) | 0.23 |
|  | PFOA (+) | 0.50 | PFOA (+) | 0.17 | PFOA (+) | 0.28 |
| Change  2022-2016 | PFOS (-) | 0.36 | PFOS (-) | 1.0 | PFOS (-) | 1.0 |
|  | PFNA (-) | 0.39 | PFNA (+) | 0.41 | PFNA (+) | 0.15 |
|  | ETU (-) | 0.25 | ETU (+) | 0.14 | ETU (+) | 0.21 |
|  | Glyphosate (+) | 0.13 | Glyphosate (+) | 0.06 | Glyphosate (+) | 0.21 |
|  | 2,4D (+) | 0.37 | 2,4D (+) | 0.22 | 2,4D (+) | 0.17 |
